# Inhaled Corticosteroids for Outpatients with Covid-19: A Meta-Analysis

**DOI:** 10.1101/2021.11.04.21265945

**Authors:** Todd C. Lee, Émilie Bortolussi-Courval, Sara Belga, Nick Daneman, Adrienne K. Chan, Ryan Hanula, Nicole Ezer, Emily G. McDonald

**Affiliations:** Division of Infectious Diseases, Department of Medicine, McGill University Health Centre, Montréal, Canada; Clinical Practice Assessment Unit, Department of Medicine, McGill University Health Centre, Montréal, Canada; Division of Experimental Medicine, Department of Medicine, McGill University, Montréal, Canada; Division of Infectious Diseases, Department of Medicine, University of British Columbia, Vancouver, Canada; Division of Infectious Diseases, Department of Medicine, Sunnybrook Health Sciences Centre, Toronto, Canada; Division of Respirology, Department of Medicine, McGill University Health Centre, Montréal, Canada; Division of General Internal Medicine, Department of Medicine, McGill University Health Centre, Montréal, Canada

**Author notes:** **Corresponding author:** Todd C. Lee MD MPH, 1001 Decarie Blvd E5-1820, Montreal, QC, H4A3S1. For the purposes of authorship, these authors share equal credit (Nicole Ezer, Emily G. McDonald).

## Abstract

The role of inhaled corticosteroids for outpatient COVID-19 is evolving. We meta-analyzed reported clinical trials and estimated probability of any effect and number needed to treat of 50 or 20 for symptom resolution by day 14 [100%, 99.8%, 93.1%] and hospitalization [89.3%, 72.9%, 26.7%] respectively.

## Introduction

Inhaled corticosteroids, in particular budesonide, have received substantial interest as inexpensive and safe treatments for non-hospitalized patients presenting with symptomatic SARS-CoV-2 infections following two open label randomized controlled trials (RCTs). STOIC (Steroids in COVID-19, n=146) [1] was the first to report budesonide was effective at improving time to recovery and reducing the composite outcome of urgent care, emergency room visits, and hospitalization. Shortly thereafter, the PRINCIPLE trial (Platform Randomized Trial of Treatments in the Community for Epidemic and Pandemic Illnesses, n=1719 concurrent) [2] replicated the findings for time to recovery and detected a reduction in hospitalization, primarily in those older than 65. However, both STOIC and PRINCIPLE were open label. Previous work has demonstrated that, with respect to respiratory symptoms, inhaled medications can have substantial placebo effects [3]. By contrast, both the recent CONTAIN trial (Inhaled Ciclesonide for the Treatment of COVID-19 in Non-hospitalized Adults, n=203) [4] and an industry-sponsored ciclesonide trial (Covis Pharma, n=400) [5] were placebo-controlled and failed to demonstrate a benefit in time to recovery with conflicting findings on hospitalizations. To inform clinical practice, we conducted a meta-analysis of these trials to contextualize the totality of the data in terms of expected effect sizes when treating symptomatic outpatients with COVID-19.

## Methods

We used the secondary outcome result of complete resolution of symptoms by Day 14 which was conserved between all four completed and reported inhaled corticosteroid randomized controlled trials: STOIC [1], PRINCIPLE [2], CONTAIN [4], and Covis Pharma [5]. We also compared the outcome of hospitalization; for STOIC, only the composite data was reported. Using STATA version 17 and the metan command, we performed a random effects meta-analysis for these outcomes stratified by the presence or absence of placebo control with a pooled overall effect estimate. With the estimates for risk ratio (RR) and the accompanying 95% confidence interval, we calculated the probability of any benefit (RR>1 for symptom resolution, RR<1 for hospitalization) as well as for a 5% (NNT of 20) and 2% (NNT 50) absolute difference based on the overall control event rates (29.3% for symptomatic improvement; 10.2% for hospitalization) integrating the area under the probability density curves [6]. We repeated the above with a fixed effects model as a sensitivity analysis.

## Results

The four trials included a total of 2317 analyzed patients and are summarized in Table 1. The average age in patients enrolled in the STOIC, CONTAIN and Covis Pharma studies were similar ranging from 37 to 45, whereas the average age of patients in the PRINCIPLE trial was much higher at 64. The pooled relative risk and 95% confidence intervals for complete symptom resolution by day 14 and hospitalization are also reported in Table 1. As hypothesized, the effect size for symptomatic improvement was larger in the open-label trials (RR 1.39; 95%CI 1.22-1.58) than in the placebo-controlled studies (RR 1.15; 95%CI 0.95-1.38). However, even the placebo-controlled studies suggest a 92.5% probability of any benefit and a 78.1% probability of an NNT ≤50. There was little heterogeneity, thus the random and fixed effects models were very similar. Whereas the open label studies individually suggest a high probability of reduction in hospitalization (RR 0.44; 95%CI 0.12-1.70; 89.3% probability of any effect), the placebo-controlled estimate was more modest (RR 0.90; 95% CI 0.22-3.71; 54.7% probability of any effect). There was moderate heterogeneity, with the fixed effect model showing higher probability of any effect (99.0% vs 89.3%) with similar probability of an NNT ≤50 (78.2% vs. 72.9%) a much lower probability of an NNT ≤20 (0.7% vs. 26.7%).

**Table 1.** Trial descriptions and Meta-Analysis Results.

| Study | Timing and Primary Outcome | Total Patients | Average Age | Male (%) | Comorbidities | Symptom Free by Day 14 |  | Hospitalized |  |
| --- | --- | --- | --- | --- | --- | --- | --- | --- | --- |
|  |  |  |  |  |  | Drug | Control | Drug | Control |
| CONTAIN (Placebo) | ≤6 days of symptoms<br>Resolution of cough, dyspnea, and fever day 7 | 203 | 37 | 46.3% | 20.2% Overall<br>5.9% HTN<br>2.5% Diabetes<br>0.5% CAD | 57/105 | 44/98 | 6/105 | 3/98 |
| Covis Pharma (Placebo) | ≤72h of test<br>Time to symptom free | 400 | 43 | 44.8% | 22% HTN<br>7.5% Diabetes | 81/197 | 76/203 | 3/197 | 7/203 |
| STOIC (Open label) | ≤7d of symptoms<br>Covid-19 urgent visits | 139 | 45 | 42.4% | Median of 1<br>8.4% CAD<br>5% Diabetes | 63/70 | 48/69 | 2/70 | 11/69 |
| PRINCIPLE (Open label) | ≤14d of symptoms<br>Covid-19 related hospitalization or death | 1719 (concurrent) | 64 | 48.5% | 80% (median of 1)<br>43-46% HTN<br>20-23% Diabetes<br>15-17% CAD | 251/781 | 173/794 | 72/787 | 98/799 |
| Analysis |  | Pooled Risk Ratio |  | Probability Any Benefit |  | Probability NNT ≤50 |  | Probability NNT ≤20 |  |
|  |  | Random | Fixed | Random | Fixed | Random | Fixed | Random | Fixed |
| Symptom Free Day 14 (I<sup>2</sup> 30%) |  | 1.29 (1.14-1.47) | 1.31 (1.18-1.45) | 100% | 100% | 99.8% | 100% | 93.1% | 98.2% |
| Placebo-controlled (I<sup>2</sup> 0%) |  | 1.15 (0.95-1.38) | 1.15 (0.95-1.38) | 92.5% | 92.7% | 78.1% | 78.2% | 42.6% | 43.4% |
| Open label (I<sup>2</sup> 11.9%) |  | 1.39 (1.22-1.58) | 1.39 (1.23-1.56) | 100% | 100% | 100% | 100% | 99.3% | 99.8% |
| Hospitalization – Overall (I<sup>2</sup> 49.2%) |  | 0.64 (0.31-1.29) | 0.72 (0.55-0.95) | 89.3% | 99.0% | 72.9% | 78.2% | 26.7% | 0.7% |
| Placebo-controlled (I<sup>2</sup> 54.4%) |  | 0.90 (0.22-3.71) | 0.90 (0.35-2.33) | 54.7% | 57.6% | 43.0% | 40.1% | 21.6% | 12.3% |
| Open label (I<sup>2</sup> 71.3%) |  | 0.44 (0.12-1.70) | 0.71 (0.59-0.94) | 89.1% | 99.8% | 81.3% | 85.3% | 57.6% | 0.3% |

## Discussion

This is the first meta-analysis of the four completed and reported trials of outpatient inhaled corticosteroid COVID-19 treatment. Our results support the use of inhaled corticosteroids (ciclesonide or budesonide) for the resolution of symptoms at day 14 of treatment. While there is undoubtedly a placebo effect, the probability of an objective effect remains high in the placebo-controlled subgroup at 92.5% probability for any effect and 78.1% probability of an NNT of 50 or less. Overall, inclusive of any additional placebo effect, there is at least a 93.1% chance that the NNT is ≤20. With respect to hospitalization, the effect is promising, but less clear due to the large influence of the PRINCIPLE trial which included a much older population compared to the other trials combined with lack of distinction between urgent care visits and hospitalization in STOIC. While the statistical test for heterogeneity in PRINCIPLE was not significant, there was a notable and plausible difference in the subgroup of patients older than 65 (aOR 0.60; 95%CI 0.40-0.90) when compared to younger participants (aOR 1.03; 95%CI 0.59-1.80). Overall, the probability of a clinically significant effect on hospitalization (NNT ≤50) was only 72.9% (78.2% in the fixed model) and this further may be an overestimate because the pooled control event of 10.2% was driven by PRINCIPLE and STOIC. If using inhaled corticosteroids to prevent hospitalization, the yield will be higher with greater patient risk.

Our analysis is limited by the granularity of the available data. An individual patient meta-analysis accounting for age and comorbidities might produce more accurate estimates, particularly in subgroups. Furthermore, individual patient date would facilitate time to event analyses which could have increased power. Additionally, approximately two-thirds of the data is open label and subject to the placebo effect with respect to symptom reporting. There is potentially bias in urgent care or emergency room utilization due to unblinded providers being less likely to refer to urgent care when the patient was on treatment, and/or a difference in care-seeking behavior for participants. Finally, these trials were performed in different waves of the global COVID-19 pandemic. Patients and providers may have been more likely to refer patients to the emergency department early in the pandemic when less was known about the natural history of the disease. If additional placebo-controlled trials become available, it will be important to update any meta-analysis. The strength of this analysis is that we have used all the available data in combination with a probabilistic presentation allowing for determination of a variety of clinically relevant effect sizes. Inhaled corticosteroids are widely available, inexpensive in most jurisdictions, have few reported severe side effects, and are likely beneficial based on the total evidence to date.

Overall, there is an ongoing need to identify available, affordable, and effective oral or inhaled medications that can be used early in the disease to prevent COVID-19 hospitalization. It is unknown whether improving complete symptom resolution will have any meaningful impact on long term outcomes and the prevention of chronic symptoms that are now commonly reported. With respect to reduction in hospitalization, there is promise for inhaled corticosteroids, particularly in older adults; however, additional placebo controlled randomized trial evidence should still be sought to minimize bias and obtain more accurate estimates of effect size.

**Figure 1.**
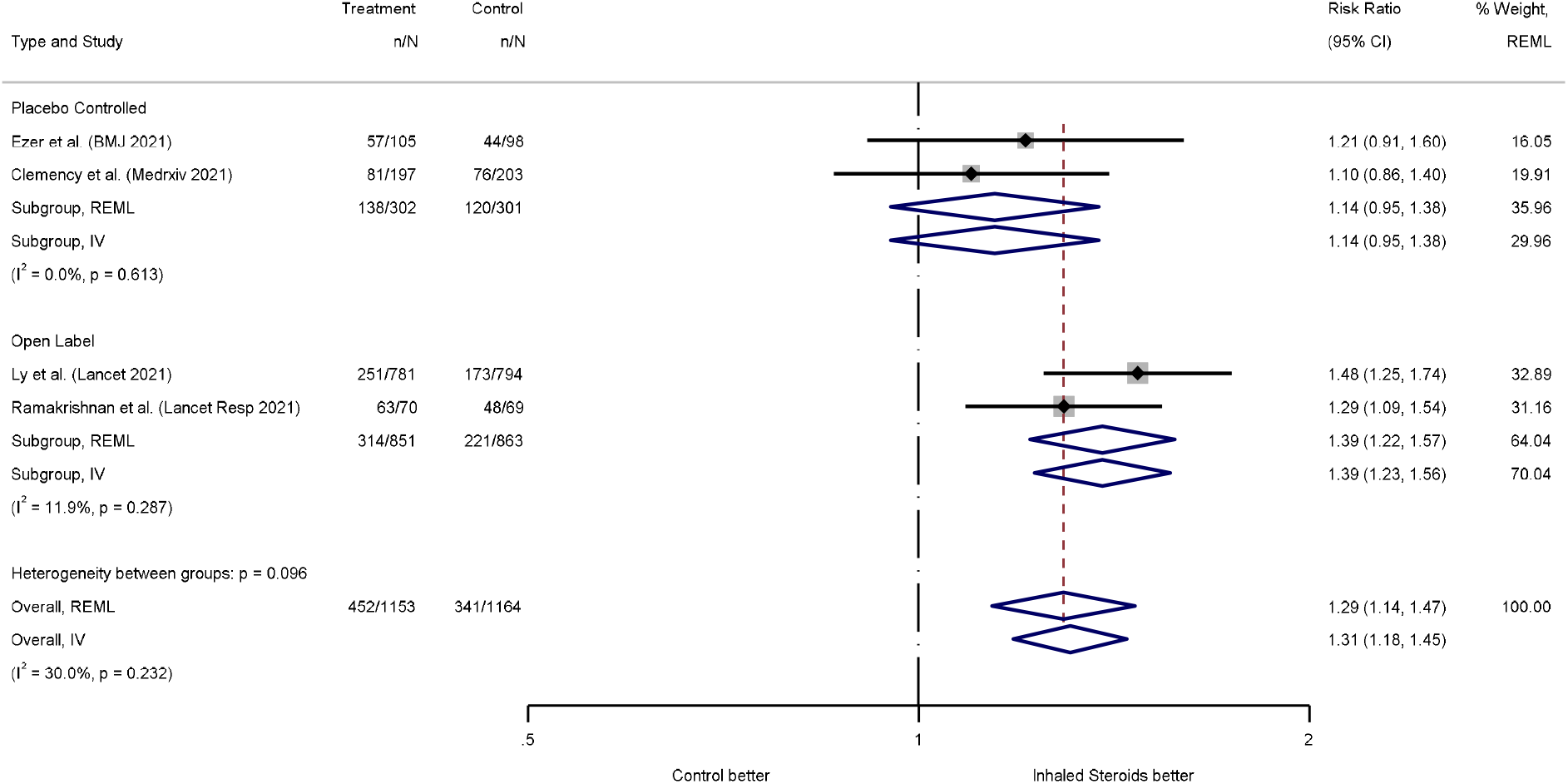
Improvement of Symptoms by Day 14.

**Figure 2.**
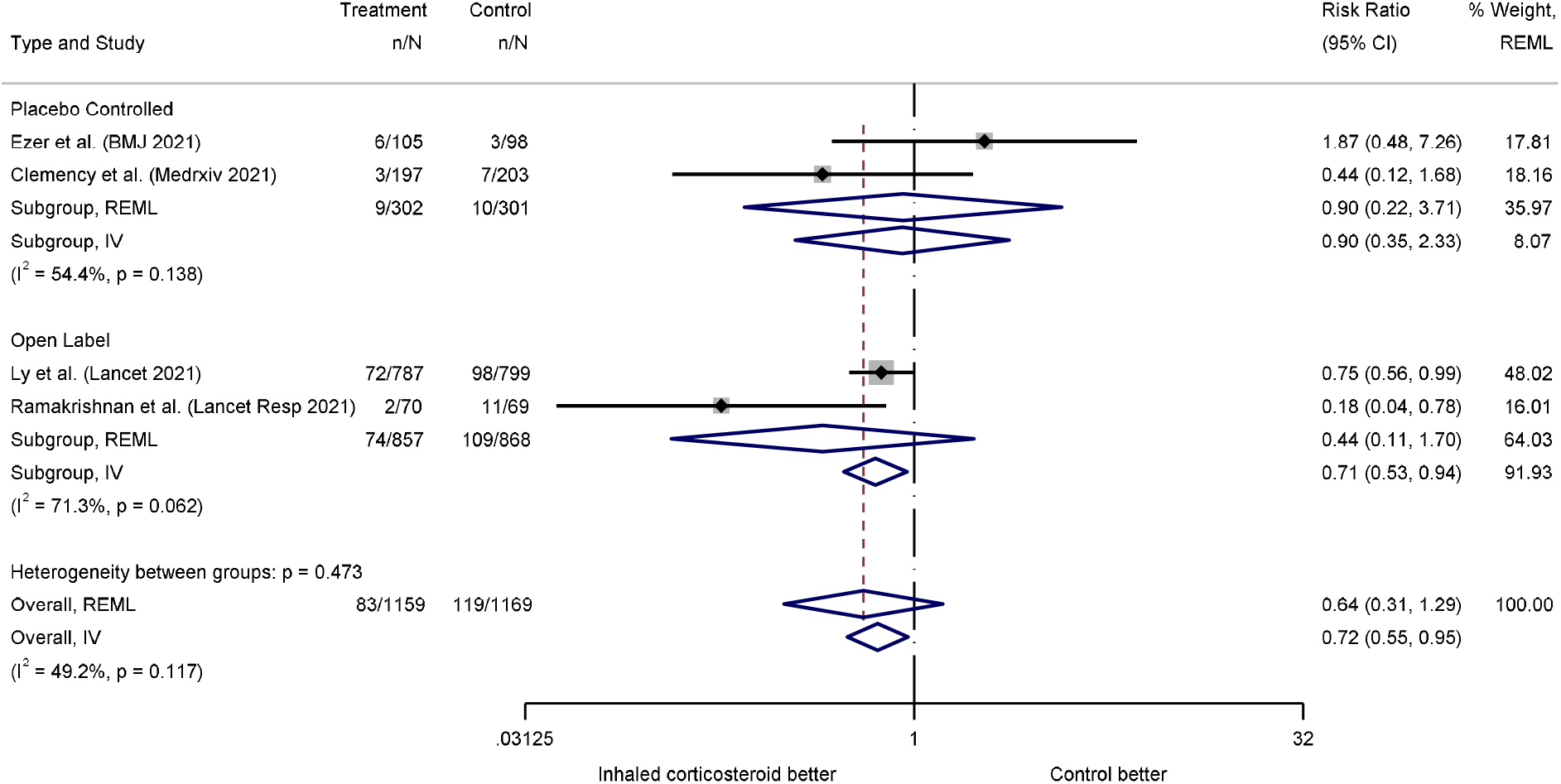
Hospitalization.

## Data Availability

All data produced in the present work are contained in the manuscript

## Acknowledgements

TCL, NE, and EGM receive research salary support from the Fonds de Recherche du Québec – Santé.

## CRediT author statement

Conceptualization - TCL, EGM; Methodology - TCL, EGM; Validation - TCL; Formal Analysis - TCL; Investigation - All authors; Resources - TCL; Data Curation - TCL, NE, EGM; Writing - Original Draft - TCL, ÉBC, RH, EGM; Writing - Review and Editing - All authors; Visualization TCL, EGM

## Data Sharing

Statistical code available on request from Dr. Lee

